# Lifespan of COVID-19 living guideline recommendations: a survival analysis

**DOI:** 10.1101/2023.06.08.23291123

**Authors:** Emma McFarlane, Toby Mercer, Steve Sharp, Debra Hunter, Kate Kelley, Fiona Glen, Maria Majeed

**Affiliations:** National Institute for Health and Care Excellence, Manchester, UK

**Keywords:** Living guidelines, COVID-19, lifespan of recommendations, updating guidelines

## Abstract

**Background:** NICE has maintained a portfolio of COVID-19 living guidelines since March 2020. Recommendations within these living guidelines are subject to continuous surveillance and updates in response to triggers. However, the lifespan of individual living guideline recommendations and features that may impact on whether a recommendation becomes out of date sooner, is unknown.

**Objectives:** This study aimed to describe the length of time NICE COVID-19 living guideline recommendations have remained valid.

**Methods:** All guidelines within NICE’s COVID-19 portfolio were included to determine the lifespan of living guideline recommendations. Data were collected on all recommendations that had been developed, undergone surveillance or updated between 1 March 2020 and 31 August 2022. Information on initial publication date, decision to update, and update publication date was extracted. Updates were labelled as major changes in evidence synthesis or minor changes without a substantial change in evidence base. Any recommendation that had not been updated or withdrawn was censored. Survival analysis (Kaplan-Meier Curve) was carried out to determine the lifespan of recommendations.

**Results:** Overall, 26 COVID-19 living guidelines and 1182 recommendations were included in the analysis. Living recommendations had median survival time of 739 days (IQR: 332, 781). Based on recommendation type, intervention recommendations had a shorter survival time (354 days, IQR 312, 775) compared to diagnosis (368 days, IQR: 328, 795), patient experience (733 days, IQR: 345, 795) and service delivery (739 days, IQR: 643, 781). Within intervention type, pharmacological recommendations had shortest survival time versus non-pharmacological recommendations [335 days (IQR: 161, 775) vs 775 days (IQR: 354, 775)]. Updates were published an average of 29.12 days following a surveillance decision.

**Conclusion:** Within living guidelines, some recommendations need to be updated sooner than others. This study outlines the value of a flexible responsive approach to surveillance within the living mode according to pace of change and expectation of update triggers.

**What is new?:** *Key findings:* Within the context of a living guideline, some recommendations will become out of date sooner than others.

*What this adds to what was known?:* This study supports the concept of prioritising recommendations within a guideline to be living.

*What is the implication and what should change now?:* Guideline developers should consider which recommendations within a living guideline would have the most value in being maintained as living to optimise resources.

## Introduction

Living guidelines provide continually updated advice based on the best available evidence [Akl et al 2017]. Living guidelines are considered to be most useful in high-priority clinical areas where there is uncertainty and a high likelihood of emerging new evidence [Cheyne et al 2023]. This approach has been useful in the context of the COVID pandemic as creating living guidelines on managing COVID-19 has enabled guideline developers to update recommendations quickly in response to dynamic changes in the evidence base.

To achieve living guidelines, surveillance is conducted at frequent intervals and evidence or system intelligence is rapidly considered for its impact on recommendations [Navarro et al 2023]. Updates to recommendations are undertaken as soon as possible after a trigger indicates the need to update a guideline. Updates to recommendations may be in response to several triggers [NICE manual 2022]:

- publication of a study that is directly relevant to the guidance and has the potential to affect existing recommendations
- substantial changes in policy or legislation (an example includes changes to the UK physical activity guidelines by the Chief Medical Office)
- withdrawal of a drug from the market, or a clinically significant drug safety update from the Medicines and Healthcare products Regulatory Authority or the Commission on Human Medicines.
- real-world data indicating a change in the disease that significantly changes the hospitalisation or mortality rate

In a living guideline, the unit of update is generally individual recommendations meaning these can be updated as relevant new evidence emerges, without the need to wait for the entire guideline to be revised [Akl et al 2021].

A previous survival analysis of a cohort of published NICE clinical guidelines found that the median lifespan of guidelines was 60 months [Alderson et al 2014]. Although it is recognised that individual recommendations within a living guideline will be faster moving than others, it is not known how long individual living recommendations remain valid and whether there are particular factors that affect their lifespan.

This study aimed to describe the length of time NICE COVID-19 living guideline recommendations have remained valid.

## Materials and methods

All NICE guidelines on COVID-19 published between 1 March 2020 and 22 August 2022 were included. The guidelines were all maintained as living guidelines and therefore eligible for inclusion. The living approach included continuous surveillance for new emerging evidence and conducting updates as soon as possible after a trigger for update emerged. Overall, 26 guidelines related to COVID-19 were included covering service delivery, modifications for other conditions and specific management of COVID-19 and its complications (see Appendix 1 for the full list). As these were living guidelines, the unit of surveillance and updates was at the recommendation level.

### Data Extraction

Data was extracted at individual recommendation level for each guideline into an Excel spreadsheet. Data was extracted by one reviewer and quality assured by a second reviewer. Disagreements were resolved through discussion with a third reviewer. Data extraction was agreed a priori and recorded in a protocol for reviewers to follow. Key dates were logged including date of first publication, surveillance trigger date and date of update publishing. Descriptive data was also collated per recommendation including guideline number, guideline title, recommendation number, section number and title, question type, topic area and underpinning evidence base. The 26 guidelines were published in one of two formats, html on the NICE website or in [MAGICapp](https://app.magicapp.org/#/guidelines). For the html guidelines, the recommendation number allocated in the guideline was used as a unique ID whereas the MAGICapp recommendations were given sequential recommendation numbers manually as an ID (NICE guidelines NG191 and NG188). Each recommendation was categorised to one of the following review types: intervention, diagnosis, prognosis, service delivery, patient experience, aetiology or health inequalities. Intervention recommendations were allocated an additional sub-category of either pharmacological or non-pharmacological. Interventions that were related to therapeutics or medicines for management of symptoms and disease were allocated to the pharmacological subcategory. Non-pharmacological interventions included management approaches such as respiratory support and rehabilitation. Recommendations were also categorised into one of four types of evidence that was judged to underpin the recommendation including quantitative, qualitative, mixed evidence (defined as both quantitative and qualitative) and committee consensus. The category of committee consensus was used where no references were related to the recommendation. For the other categories, the evidence review used to inform the recommendation was used to judge the evidence type.

Updates were also categorised as minor or major. Minor changes included adding or deleting hyperlinks, amending text for clarity, deleting outdated information, updating information taken from other guidelines and changing supplementary information (such as referring to an updated national policy from an external source). Major changes were categorised as a change in evidence base including conducting a new meta-analysis or updating an existing meta-analysis with new studies or outcomes that resulted in a change or no change in the recommendation wording or intent. Additionally, if a recommendation or whole guideline was withdrawn or incorporated into another guideline, this was also considered a major change.

A decision to update was recorded if a recommendation had undergone surveillance and the decision had been reached to ‘update’. In this scenario, a date of update publishing was added to the recommendation data. The subsequent publication of the updated recommendation was then logged on a new row of the spreadsheet as another datapoint. The updated recommendation was further assessed for surveillance triggers until withdrawn or censored for survival time.

If a recommendation had undergone surveillance but no update, the decision logged was ‘no update’. If a recommendation had not undergone surveillance, the decision to update was coded as ‘no surveillance’ and censored on 31 August 2022 which was the cut-off date for the study.

### Data Analysis

Survival time per recommendation was calculated in weeks from publication date to the date of update publishing, withdrawal, or censored date, in cases where there had not been any surveillance.

#### Subgroup Analyses and Rationale

Subgroup analyses were conducted based on distinct review types, encompassing intervention, diagnosis, prognosis, service delivery, patient experience, aetiology, and health inequalities. The primary objective of this analysis was to elucidate disparities in recommendation lifespans across diverse review types, offering valuable insights into areas that may necessitate more frequent updates or heightened surveillance.

A further subgroup analysis was carried out within the intervention recommendations category, contrasting pharmacological and non-pharmacological interventions. This evaluation aimed to investigate potential variances in the lifespan of recommendations pertaining to drug therapies and other management approaches, such as respiratory support and rehabilitation.

An additional subgroup analysis compared the lifespans of recommendations based on the underpinning evidence base, which included quantitative, qualitative, mixed evidence, and committee consensus. This examination aimed to assess the impact of the type of evidence on the recommendation lifespan and ascertain whether specific types of evidence may contribute to more frequent updates or shorter lifespans.

Lastly, a subgroup analysis compared the lifespan of recommendations when minor and major updates was conducted. This investigation sought to determine whether the type of update (minor changes or significant changes to the evidence base) influenced the lifespan of living guideline recommendations.

### Statistics

Stata version 13 software was used to calculate median survival times, interquartile ranges, and to plot Kaplan-Meier survival estimate curves for all data and for each subgroup analysis. The median survival time was taken as the timepoint at which the survival probability dropped to 0.5 or 50%.

Microsoft Excel was used to calculate the duration of time in days between the surveillance trigger dates to the dates of an update publishing or withdrawal of a recommendation. Mean values were then calculated in Excel for all data and for each subgroup analysis.

## Results

The results are shown in Table 1. In total, 1182 recommendations were included from 26 COVID-19 living guidelines. The median survival time of living recommendations was 739 days IQR: 332, 781 (see chart 1). Subgroup analyses indicated that prognostic recommendations had the shortest lifespan compared with service delivery recommendations which had the longest lifespan (medians of 328 versus 739 days IQR 643, 781 respectively) (see chart 2). However, only 10 recommendations were categorised as prognostic so there is considerable uncertainty around this result. Intervention recommendations were found to have the second shortest lifespan at 354 days IQR 312, 775 compared to diagnosis (368 days IQR: 328, 795), patient experience (733 days IQR: 345, 795) and service delivery (739 days IQR: 643, 781).

**Chart 1:**
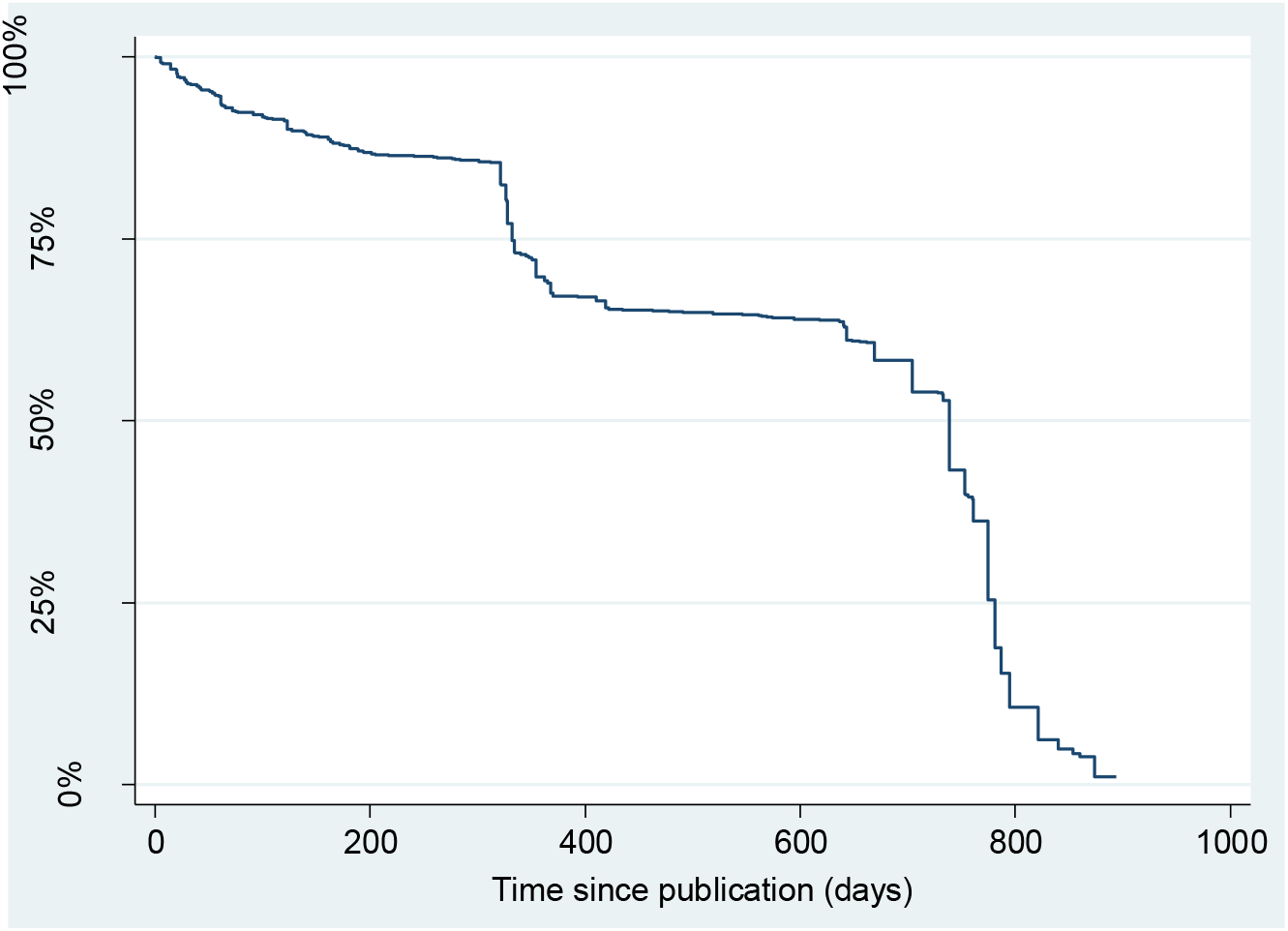
Kaplan-Meier survival curve for all recommendations.

**Chart 2:**
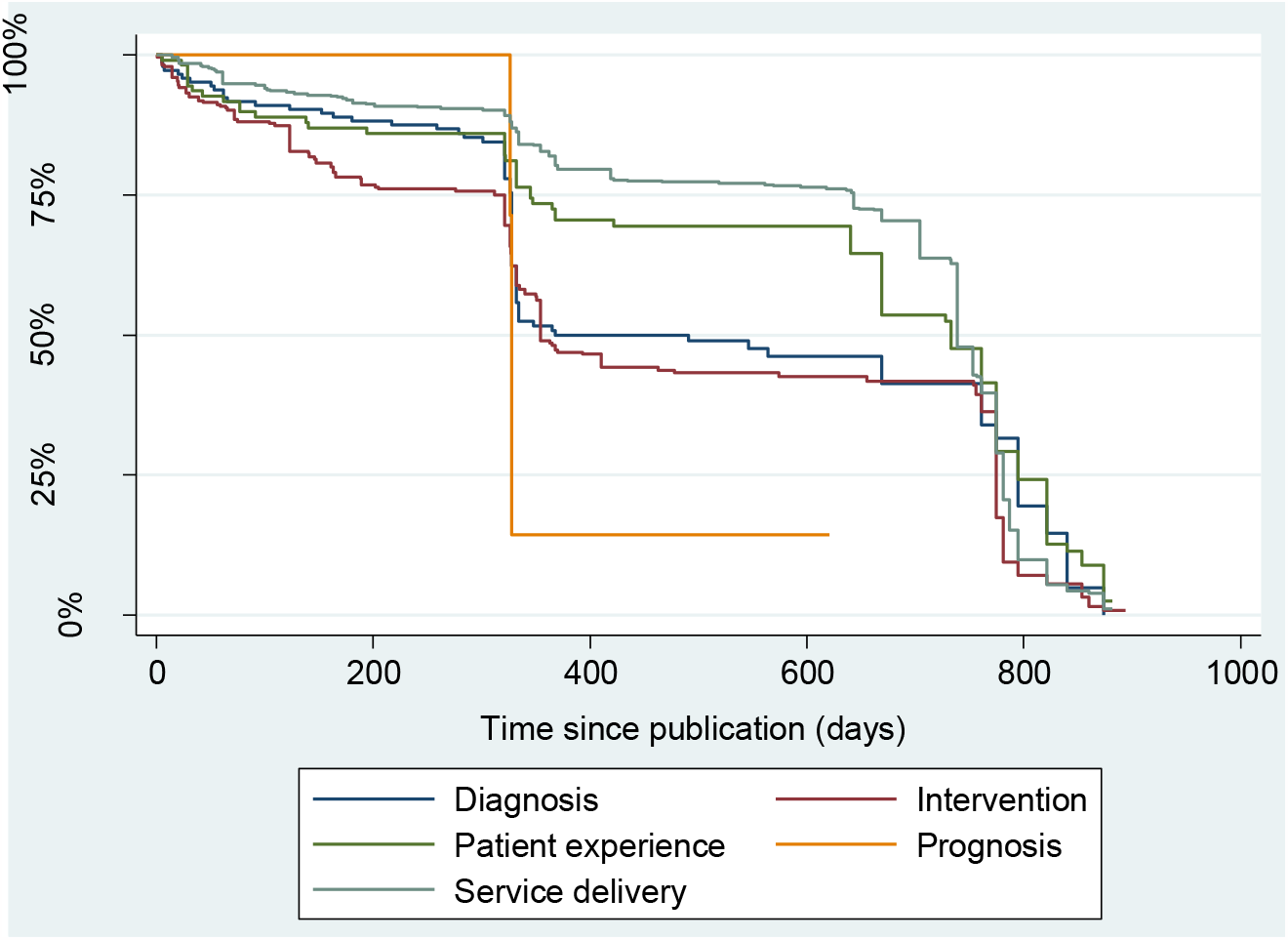
Kaplan-Meier survival curves for the recommendation types.

**Table 1:** Results for all categories of analysis.

| Category of analysis | Number of recommendations that were published<sup>a</sup> | Total number of recommendation days analysed<sup>b</sup> | Incidence rate of update or withdrawal per day | Mean days from first surveillance trigger to date of update, publishing, or withdrawal | Survival time |  |  |
| --- | --- | --- | --- | --- | --- | --- | --- |
|  |  |  |  |  | 25<sup>th</sup> percentile | 50<sup>th</sup> percentile (median average) | 75<sup>th</sup> percentile |
| Total for all recommendations | 1182 | 616293 | 0.0015463 | 29.12 | 332 | 739 | 781 |
| **By recommendation type** |  |  |  |  |  |  |  |
| Diagnostic | 146 | 58418 | 0.0014721 | 42.41 | 328 | 368 | 795 |
| Interventions: total | 296 | 112065 | 0.0018471 | 27.78 | 312 | 354 | 775 |
| Interventions: pharmacological | 205 | 69584 | 0.0019401 | 23.40 | 161 | 335 | 775 |
| Interventions: non-pharmacological | 80 | 39748 | 0.0015850 | 23.40 | 354 | 775 | 775 |
| Interventions: pharmacological with non-pharmacological | 11 | 2733 | 0.0032931 | 26.22 | 163 | 363 | 368 |
| Patient experience | 110 | 58542 | 0.0014690 | 47.78 | 345 | 733 | 795 |
| Prognostic | 10 | 3464 | 0.0017321 | 37.33 | 326 | 328 | 328 |
| Service delivery | 620 | 383804 | 0.0014799 | 24.65 | 643 | 739 | 781 |
| **By evidence type** |  |  |  |  |  |  |  |
| Consensus | 1074 | 574791 | 0.0015275 | 29.28 | 334 | 739 | 781 |
| Mixed | 48 | 28587 | 0.0016441 | 20.66 | 463 | 669 | 775 |
| Quantitative | 60 | 12915 | 0.0021680 | 38.32 | 123 | 321 | c |
| **By evidence type for the intervention recommendations alone** |  |  |  |  |  |  |  |
| Consensus | 218 | 85390 | 0.0018620 | 27.45 | 321 | 354 | 775 |
| Mixed | 20 | 14276 | 0.0014010 | 15.65 | 775 | 775 | 775 |
| Quantitative | 58 | 12399 | 0.0022582 | 38.32 | 123 | 205 | c |
| **By major or minor update** |  |  |  |  |  |  |  |
| Major | 791 | 490329 | 0.0016132 | 28.34 | 363 | 739 | 781 |
| Minor | 163 | 30053 | 0.0053905 | 32.92 | 33 | 100 | 328 |
| Major or minor<sup>d</sup> | 954 | 520382 | 0.0018313 | 29.12 | 328 | 704 | 775 |
| a) This includes the number of recommendations that were published and updated. In other words, when a recommendation is updated, we consider the update to be a different and new recommendation.<br>b) This may also be called 'recommendation days at risk'<br>c) The 75<sup>th</sup> percentile is not calculable because fewer than 75% of the recommendations in this category were updated or withdrawn by the censor date of 31/8/22<br>d) These values are different compared with the total for all recommendations because 228 recommendations were not updated by the censor date of 31/8/22 |  |  |  |  |  |  |  |

Within the intervention recommendations category, pharmacological recommendations had the shortest survival time compared with non-pharmacological recommendations (medians of 335 days versus 775 days respectively) (see chart 3).

**Chart 3:**
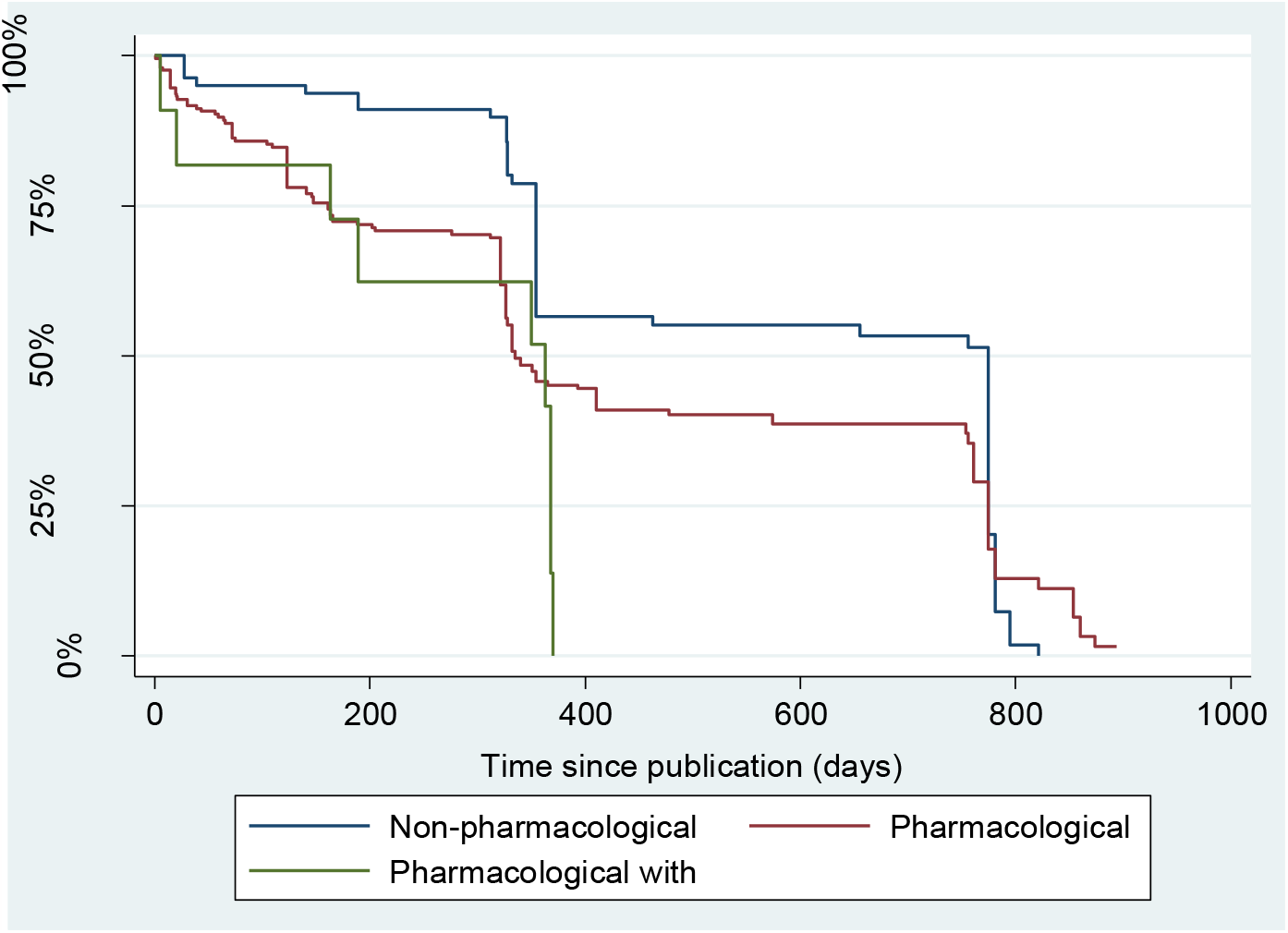
Kaplan-Meier survival curves for intervention recommendation subgroups.

Subgroup analysis comparing the lifespan when considering the underpinning evidence base for the recommendations indicated that quantitative recommendations had the shortest lifespan compared with consensus recommendations (medians of 321 versus 739 days respectively) (see chart 4).

**Chart 4:**
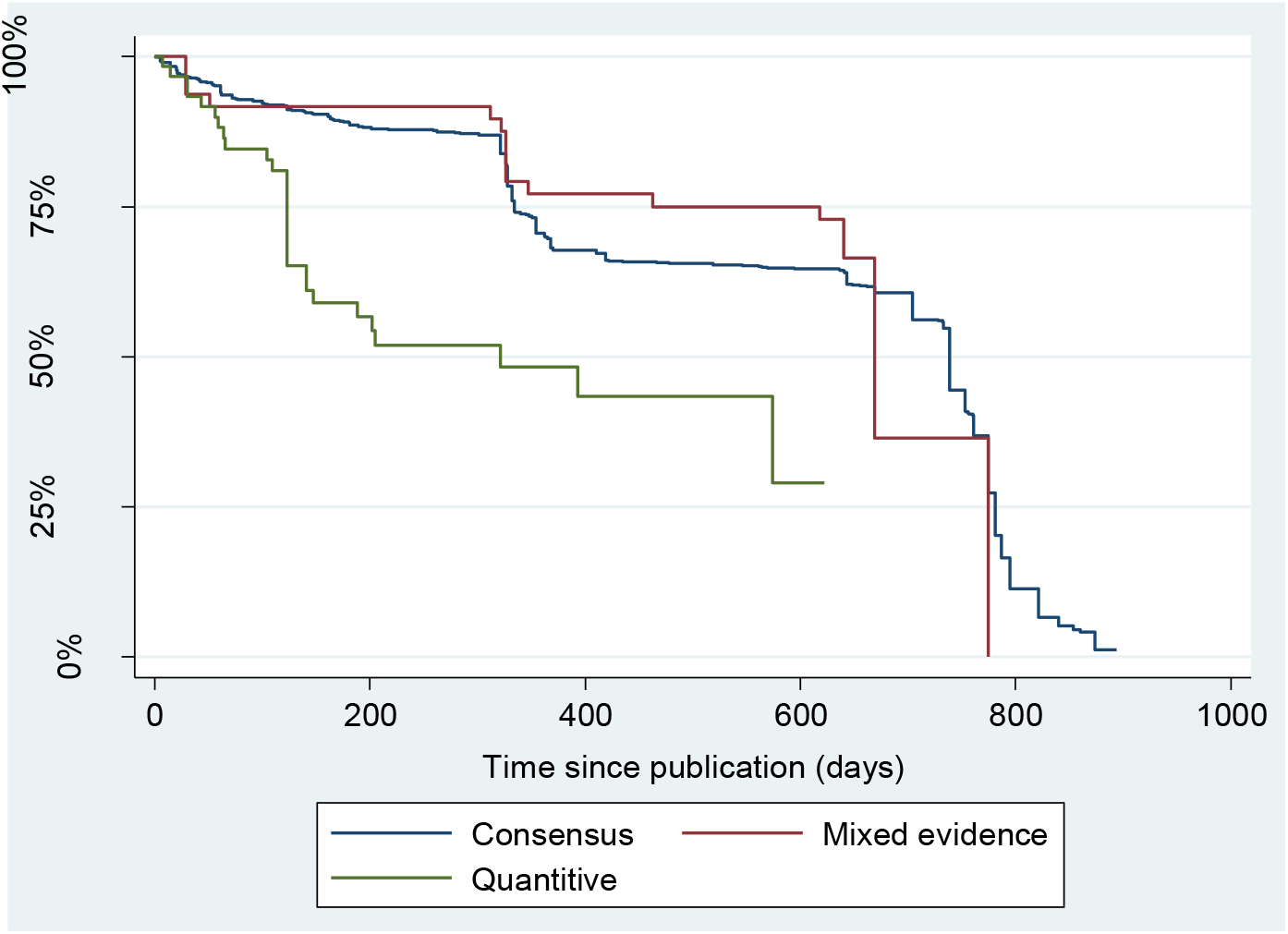
Kaplan-Meier survival curve for recommendation evidence types.

A comparison of update types showed that minor updates have a much shorter lifespan than recommendations with major updates (medians of 100 versus 739 days respectively) (see chart 6).

**Chart 5:**
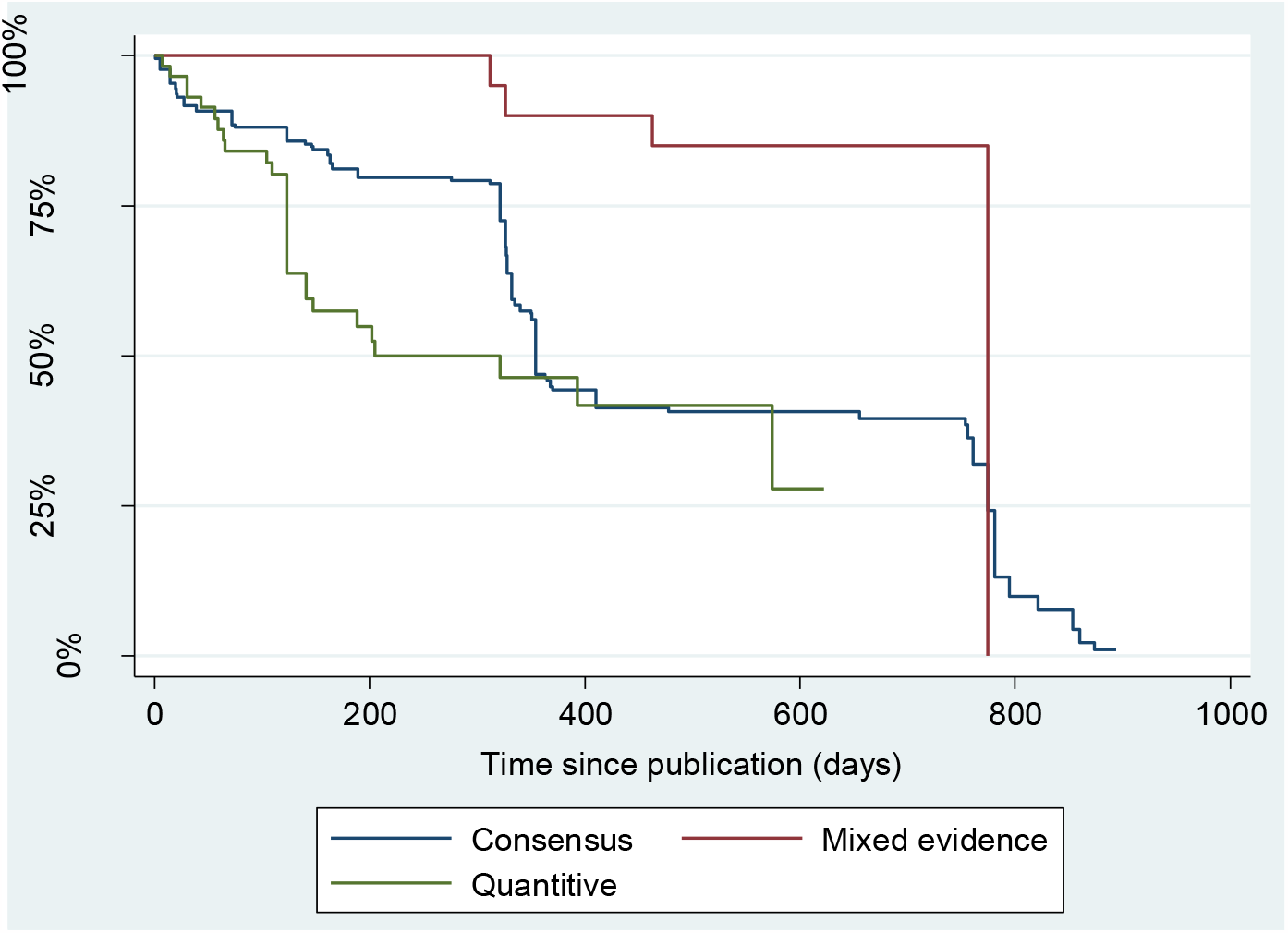
Kaplan-Meier survival curve for evidence type for the intervention recommendations alone.

**Chart 6:**
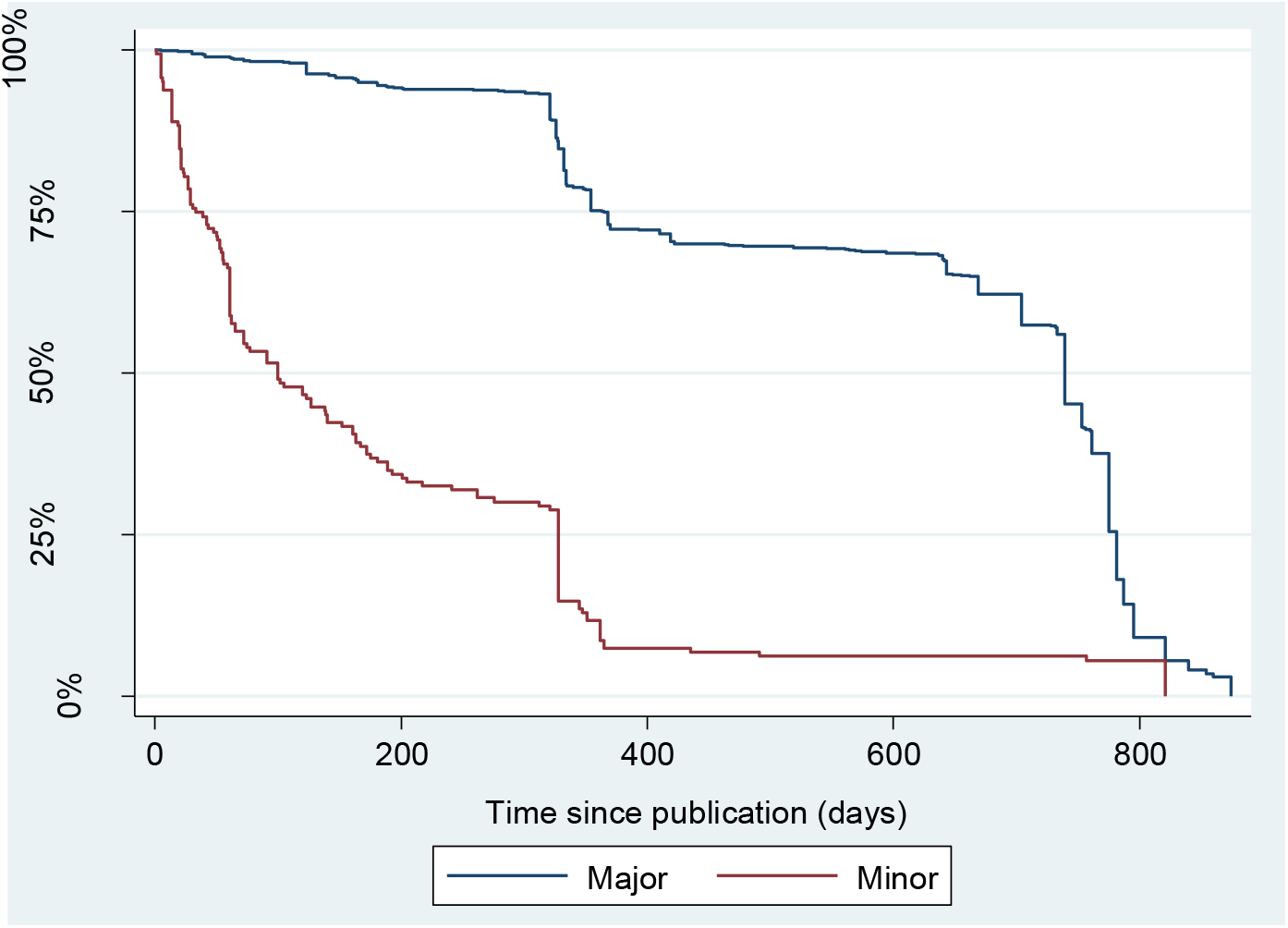
Kaplan-Meier survival curve for major or minor update.

The mean duration of time from a surveillance decision to publishing an update was 29.12 days.

Mixed evidence had the shortest surveillance trigger date to date of publishing and quantitative evidence had the longest (means of 15.65 versus 38.32 days respectively).

## Discussion

To our knowledge, this is the first study investigating the lifespan of living guideline recommendations. The results of the study demonstrated that the lifespan of recommendations differs depending on characteristics such as review type or the underpinning evidence that informed the recommendation.

The median survival time of living recommendations was 739 days which, although shorter than has been reported in previous analyses of full guidelines which has been around 5 years, is possibly longer than anticipated for living guideline recommendations (Shekelle et al 2001 and Alderson et al 2014).

In this study, it was observed that major updates exhibited a significantly longer survival time in comparison to minor updates (781 days versus 100 days, respectively). This may be attributed to the fact that major updates rely on new evidence emerging that requires an update to an evidence review whereas, in this study, minor updates tended to be editorial in nature and often prompted by user feedback following the publication of a new recommendation.

Recent studies reporting on methods for living guidelines discuss the importance of prioritising recommendations within a guideline to be living as not all recommendations are expected to require updates at the same pace (Fraile Navarro et al, 2023 and El Mikati et al, 2022). Guideline developers may want to consider the recommendations within a living guideline and the factors that could impact on how quickly they could become out of date as part of prioritisation for living mode. Sub-topic variations within living sections of a guideline may also warrant differing intensities of surveillance, such as by varying the frequency of searching and screening. The value of utilising a flexible responsive approach to surveillance within the living mode enables guideline developers to allocate resources according to pace of change and expectation of update triggers (Sharp et al. 2023).

The mean duration of time from a surveillance decision to publishing an update was 29.12 days This short timeframe is likely to have been driven by a large proportion of major updates than minor updates (791 versus 163). In subgroup analysis, shorter mean duration from surveillance to update was noted in recommendations on service delivery (24.65 days) and intervention (27.78 days) compared to recommendations on diagnosis (42.4 days), prognosis (37.3 days) and patient experience (47.7 days). In the total sample, there were 620 recommendations on service delivery and a major update was common for service delivery recommendations as withdrawal of these recommendations. Likewise, there were 296 recommendations on interventions and minor updates were more commonly observed for intervention recommendations. Both service delivery and intervention recommendations constituted a bigger part of our sample and hence may explain the overall shorter duration from surveillance decision to update publishing in the data. Other factors likely contributing to the rapid updates was the infrastructure used to deliver the COVID-19 living guidelines including reuse of data from other organisations and a rolling expert panel that could be convened when needed. Integrating stages of guideline development, surveillance and updating into a single working model has been reported by other guideline organisations and was shown to enhance the speed of the workflow (Tendal et al 2021).

In this study, a number of subgroup analyses were conducted with the aim of understanding what factors might drive how quickly recommendations become out of date and therefore, benefit various stakeholders involved in the development and implementation of living guidelines. By understanding the differences in the lifespan of living guideline recommendations across various subgroups, guideline developers can efficiently allocate resources by concentrating on areas that require more frequent updates or surveillance.

A strength of this study is the inclusion of a portfolio of guidelines that had been maintained as living guidelines for 2 years or more. The study is timely and relevant, as it addresses the lifespan of living guidelines in the context of the rapidly changing COVID-19 pandemic. We performed a comprehensive data extraction approach by extracting data at the individual recommendation level for each guideline, ensuring a detailed analysis. The inclusion of major and minor updates reflected the range of update triggers arising in living guideline surveillance. The study considered different types of recommendations, such as intervention, diagnosis, prognosis, and service delivery, which increases its comprehensiveness and generalisability. We performed Kaplan-Meier survival estimate curves and calculated quartiles of survival time, which provides robust and reliable results and performed subgroup analyses to explore the influence of different factors on the lifespan of living guideline recommendations.

Nonetheless, the findings of this study should be considered alongside the following limitations. Firstly, as the recommendations included in this study were all derived from COVID-19 guidelines, this limits the generalisability of the lifespan of living guideline recommendations which may differ across disease areas. Additionally, the study lacks external validation of the results, which may affect the generalisability and applicability of the findings to other settings.

Due to the volume of guidelines and recommendations, we employed single reviewer data extraction. Although data for each guideline was extracted by one reviewer and quality assured by a second reviewer, using multiple reviewers for data extraction could have strengthened the reliability of the data. Linked to this, the categorisation of updates into major or minor changes could be subject to bias, as different people may have different interpretations of what constitutes a major or minor change. We aimed to minimise this by creating definitions for major and minor updates and including these in the study protocol.

Finally, the study does not assess the impact of updated recommendations on clinical practice or patient outcomes, limiting the understanding of the practical implications of the findings.

It is generally accepted that within the context of a living guideline, recommendations will vary in their lifespan and become out of date at different timepoints. This study supports the concept of prioritising recommendations in more dynamic areas of a guideline to have continuous living status. Guideline developers should consider which recommendations within a living guideline would have the most value in being maintained as fully living to optimise resources.

## Data Availability

All data produced in the present study are available upon reasonable request to the authors

## Appendix 1

| Guideline No. | Guideline title |
| --- | --- |
| NG159 | Critical care in adults |
| NG160 | Dialysis service delivery |
| NG161 | Delivery of systemic anticancer treatments |
| NG162 | Delivery of radiotherapy |
| NG163 | Managing symptoms (including at the end of life) in the community |
| NG164 | Haematopoietic stem cell transplantation |
| NG165 | Managing suspected or confirmed pneumonia in adults in the community |
| NG166 | Severe asthma |
| NG167 | Rheumatological autoimmune, inflammatory and metabolic bone disorders |
| NG168 | Community-based care of patients with chronic obstructive pulmonary disease |
| NG169 | Dermatological conditions treated with drugs affecting the immune response |
| NG170 | Cystic fibrosis |
| NG171 | Acute myocardial injury |
| NG172 | Gastrointestinal and liver conditions treated with drugs affecting the immune response |
| NG173 | Antibiotics for pneumonia in adults in hospital |
| NG174 | Children and young people who are immunocompromised |
| NG175 | Acute kidney injury in hospital |
| NG176 | Chronic kidney disease |
| NG177 | Interstitial lung disease |
| NG178 | Renal transplantation |
| NG179 | Arranging planned care in hospitals and diagnostic services |
| NG186 | Reducing the risk of venous thromboembolism in over 16s with COVID-19 |
| NG187 | Vitamin D |
| NG188 | Managing the long-term effects of COVID-19 |
| NG191 | Managing COVID-19 |
| NG200 | Vaccine-induced immune thrombocytopenia and thrombosis (VITT) |

